# Time-related biases in observational studies : an illustration with COVID-19 vaccination during immune checkpoint inhibitor therapy

**DOI:** 10.1101/2025.11.03.25339367

**Authors:** Elise Dumas, Paul Gougis, Lorenzo Gasparollo, Jean-Philippe Spano, Mats Julius Stensrud

## Abstract

Estimating treatment effects from observational data requires careful specification of the study timeline. Investigators must determine when eligibility is assessed, when treatment is assigned, and when follow-up begins. Seemingly minor choices at this stage can introduce subtle time-related biases and compromise valid inference. We show how this is relevant to a recent article suggesting that SARS-CoV-2 mRNA (COVID-19) vaccines increase the effectiveness of immune checkpoint inhibitors in cancer patients (Grippin *et al.*). We first describe the original study timeline and find several sources of bias arising from its specifications. These time-related biases lead to erroneous conclusions about protective treatment effects. We then explain how time-related biases can be mitigated by careful target trial emulation, and we specify and emulate a target trial for the study of Grippin *et* al. In this target trial emulation, the estimated 3-year overall survival was 45.7% in the vaccinated group and 43.8% in the unvaccinated group (difference, 1.9 percentage points [95% CI, −23.6 to 25.0]), substantially attenuating the benefit of COVID-19 vaccines found in the original analysis.

## Introduction

Causal inference methods have improved the credibility and transparency of observational analyses of treatment effects.^1, 2^ The target trial framework has made these methods accessible and help designing observational studies for causal inference.^3, 4^ Yet, even when investigators aim to emulate a target trial, subtle time-related biases arise if the study timeline is not carefully designed.^5–7^ Avoiding these biases requires explicit specification of the start of follow-up, the intervention assignment period, and the eligibility window.^8–10^ When the data contain observations from multiple calendar years, investigators must also define the calendar window for study inclusion. Broader inclusion periods tend to increase precision but can introduce bias if the underlying risk of outcomes changes over time.

A recent study by Grippin *et al.* reported clinical and biological evidence suggesting that SARS-CoV-2 mRNA (COVID-19) vaccination enhances the effectiveness of immune checkpoint inhibitors (ICIs) in patients with advanced cancer.^11^ The clinical evidence was derived from an observational analysis of patients with stage III/IV non–small cell lung cancer (NSCLC) and stage IV melanoma, which showed that individuals vaccinated within 100 days of ICI initiation had substantially lower risks of death and disease progression compared with unvaccinated individuals In the lung cancer cohort, 3-year overall survival was 55.7% in the vaccinated group and 30.8% in the unvaccinated group. If these differences were to reflect vaccination effects, they would have important implications for clinical practice. However, we hypothesize that the authors’ conclusions could have been driven by multiple subtle forms of time-related biases, which can be mitigated by a careful target trial emulation.

In this article, we use the lung cancer cohort from Grippin *et al* as a case study to illustrate how choices of design-specific time points can introduce systematic bias in observational analyses. We then specify and emulate a hypothetical target trial addressing the same clinical question and explain how it mitigates the identified time-related biases.

### Definition of design-specific time points

A valid emulation of a target trial requires specification of several time intervals: the eligibility period, the start and end of follow-up, the intervention assignment period, and the calendar window for study inclusion.^2^ Here we elaborate on these time intervals and relate them to the original study by Grippin *et al*.

#### Eligibility

The eligibility period is the time during which an individual meets all eligibility criteria. The investigators primarily choose these criteria to reflect the population of interest: *e.g.*, patients diagnosed with stage III/IV NSCLC initiating ICI treatment. In observational studies, investigators often add criteria on top of clinically relevant eligibility criteria for practical reasons. For example, in the main analysis of Grippin *et al.*, the investigators excluded patients who received a COVID-19 vaccine outside of the 100-day window surrounding the start date of an ICI treatment line. Thus, eligibility assessment requires knowledge of events happening after ICI initiation: a COVID-19 vaccination occurring more than 100 days after ICI initiation would preclude inclusion in the unvaccinated arm. The eligibility period is distinct from the time under study, which we describe next.

#### Time under study / Follow-up

The time zero -– also called the baseline time, time of study entry, or start of follow-up -– is the time when the follow-up of a patient starts. From the time zero and onwards, study endpoints are counted. In a clinical trial, time zero corresponds to the date of randomization. Because only eligible individuals are randomly assigned a treatment, the time of randomization is within the eligibility period. In contrast, in Grippin *et al.,* the study time zero was set at the start date of the first ICI treatment line for patients in the unvaccinated group, and at the start date of the ICI treatment line closest to the vaccination date for patients in the vaccinated group. Thus, the time zero was different in the two groups. It was also defined to be earlier than the eligibility period in both groups.

The end of follow-up is the time when patients stop to be followed up. It is usually defined as the minimum date between a list of potential events. In Grippin *et al.,* the end of follow-up was defined as the date of death or administrative censoring, whichever occurs first.

The period of follow-up, also called the time under study, is the period lasting from time zero until the date of last follow-up.

#### Intervention assignment

Consider a patient recruited to a randomized controlled trial. The time of intervention assignment occurs at randomization, that is, when the patient is allocated to one of the trial’s arms. The date of randomization also aligns with the start of follow-up. In observational studies, by contrast, there is no randomization and investigators usually define intervention groups based on treatments the patients actually took during a certain period around time zero. In this setting, the intervention assignment time must be carefully chosen, e.g., to match the start of follow-up and the eligibility period. Misalignments can lead to time-related biases. For example, in Grippin et al., the vaccinated group included patients who received a COVID-19 vaccine within 100 days surrounding ICI initiation, and the control group included patients who never received a COVID-19 vaccine. Thus, the intervention assignment period spanned from −100 to +100 days surrounding ICI initiation for the vaccinated group and extended over the entire follow-up period for the control group.

#### Calendar window for study inclusion

In most clinical trials, patients are enrolled over time, that is, there are staggered entries. Enrollment stops at a pre-specified calendar time or when the planned number of participants is recruited. In retrospective observational studies with longitudinal data, participants can be included over multiple calendar years, specified by the investigators. While broader calendar windows for study inclusion tend to yield more precise estimates, they also increase susceptibility to bias arising from calendar-time confounding. Grippin *et al.* included patients in the lung cancer cohort using retrospective hospital data spanning from January 2015 to September 2022. Because the COVID-19 vaccines were only available starting in December 2020, patients treated with ICI after December 2020 had a zero probability of being vaccinated; we say that positivity is violated.^12^ Had the investigators restricted study inclusion to patients diagnosed from December 2020 onward, both vaccinated and unvaccinated individuals would have been observable throughout the entire calendar period, thereby ensuring positivity and mitigating bias.

### Time-related bias that matters

In Grippin *et al.*, the intervention groups and eligibility were assigned based on information both before and after time zero. Therefore, the different study times were not aligned. Also, the time zero and the calendar window for study inclusion were defined differently for the two intervention groups. We now describe several ways in which these misalignments introduce systematic biases. We illustrate the potential for bias through examples that mimic the setting of Grippin *et al.,* but the arguments apply more generally.

#### 1. Immortal-time bias

In the main analysis of Grippin *et al.*, the time zero was not aligned with the date of intervention assignment. First, the vaccine group was assigned using information collected after time zero (receipt of a vaccine in the 100 days after ICI start date). Conceptually, individuals allocated to the vaccine group are guaranteed to be event-free (immortal) up to the time they received the vaccine. This results in the systematic underrepresentation in the vaccine group of fragile patients experiencing early deaths after ICI initiation, which makes it seem as if the intervention is protective (Figure 1A). This time-related bias is often referred to as immortal-time bias.^6, 9, 13^

**Figure 1:**
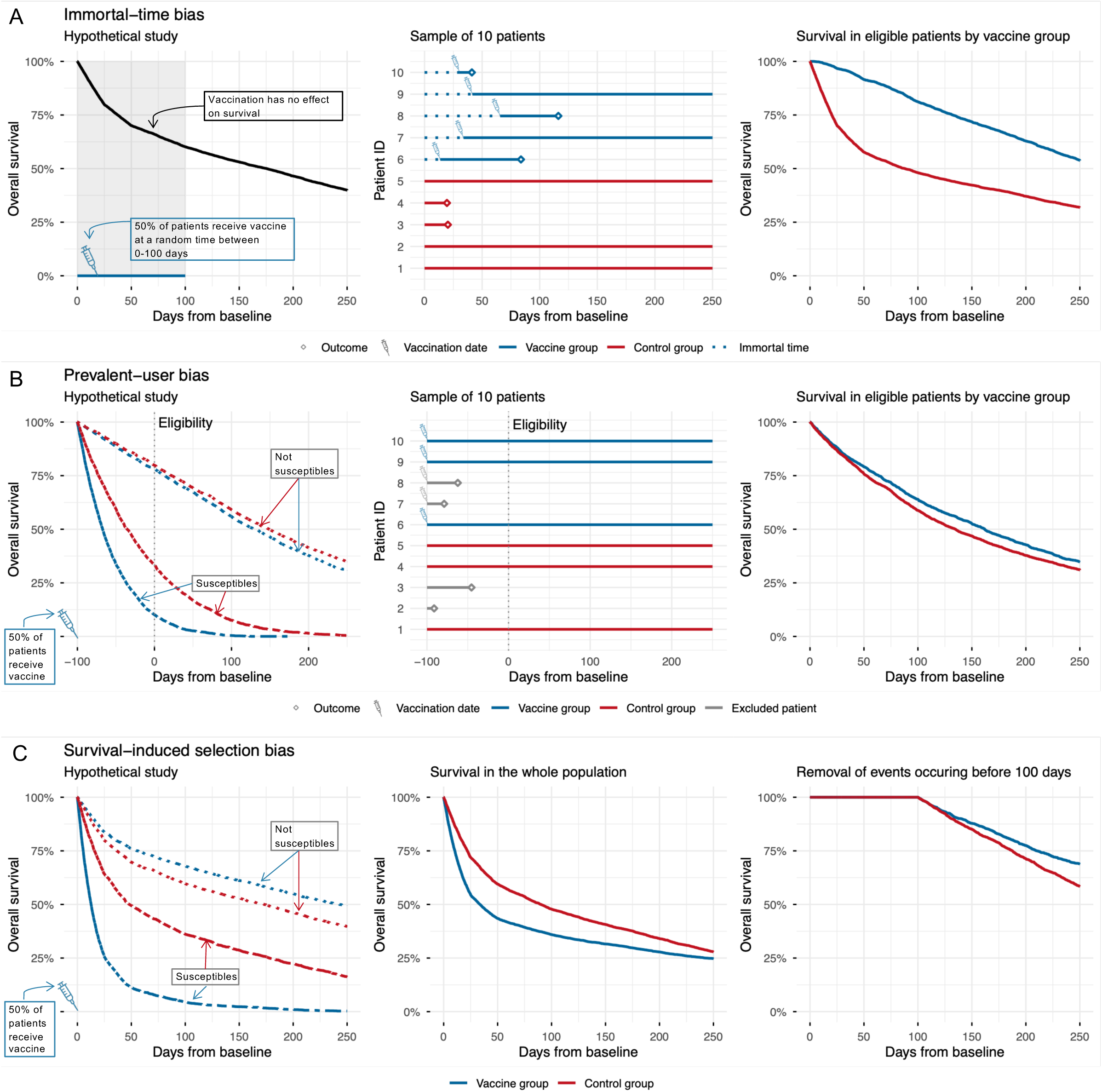
Hypothetical studies illustrating time-related biases 1, 2, and 3 in the context of Grippin et al. (A) Immortal-time bias. In this hypothetical study, vaccination has no effect on survival at any time point for any individual. Half of the patients are randomly assigned to receive a vaccine within 100 days after baseline, with vaccination dates drawn uniformly over this period. When intervention groups are defined based on receipt of vaccination within the 100 days after baseline, vaccinated patients are guaranteed to be event-free until their vaccination date. This immortal time leads to a spurious protective effect of vaccination. **(B) Prevalent-user bias.** The population consists of a mixture of susceptible and non-susceptible individuals, with susceptible patients having lower survival probabilities regardless of vaccination status. Half of the patients are randomly assigned—independently of susceptibility status—to receive vaccination 100 days before baseline. Vaccination has a harmful effect on survival in both susceptible and non-susceptible patients. However, excluding patients who died before baseline preferentially removes susceptible vaccinated individuals, resulting in vaccination appearing spuriously protective. **(C) Another form of prevalent-user bias.** The population includes susceptible and non-susceptible individuals, with lower survival among the susceptible individuals. Half of the patients are randomly assigned—independently of susceptibility status—to receive vaccination at baseline. Vaccination is harmful on average (strongly harmful in susceptible individuals and slightly protective in non-susceptible individuals). However, excluding events occurring within the first 100 days of follow-up results in vaccination appearing spuriously protective

#### 2. Prevalent-user bias

In Grippin *et al.,* patients who received vaccines in the 100 days before ICI initiation were classified into the intervention group. Therefore, at time zero, patients were already selected differently in the two vaccination groups based on previous effects of vaccines on the outcome. This is another form of time-related bias, often referred to as prevalent-user bias.^6, 14, 15^

Suppose that COVID-19 vaccines had an effect on survival because these vaccines give an immediate protection against COVID-19-related deaths in fragile (susceptible) patients. Yet the effect in patients in good shape (not susceptible) is small. Then, the most fragile patients in the unvaccinated group died of COVID-19 before they have time to initiate ICI, so that at baseline the vaccinated group includes a larger proportion of susceptible patients at baseline than the control group. Including only patients who survive long enough to initiate ICI will lead to an underestimation of the real protective effect of COVID-19 vaccination on death.

Prevalent-user bias is a form of depletion-of-susceptible –or survivor– bias.^16–18^ In contrast with immortal-time bias, it can only bias results if vaccination has an effect on the outcome, but can lead to bias in any direction and of any magnitude. For example, suppose that vaccination has a deleterious effect on survival. Then, it is possible that the effect appears protective after the exclusion of patients who are dead by the study time zero (Figure 1B).

#### 3. Postponing time zero after intervention assignment leads to another form of prevalent-user bias

Grippin *et al.* tried to address immortal-time bias by running a secondary analysis where they excluded events occurring in the first 100 days. We can interpret this analysis as postponing time zero 100 days post baseline ICI treatment for all units, *i.e.,* after intervention assignment. This procedure is also prone to prevalent-user bias, because patients susceptible to experience the outcome are unlikely to be balanced across both groups after 100 days.^6^

As an extreme illustration, suppose that COVID-19 vaccines slightly improve ICI efficacy, but also increases treatment toxicity. Increased treatment toxicity might trigger fatal adverse events in fragile (susceptible) patients, leading to vaccine increasing mortality on average. Then, at 100 days post ICI initiation, patients with the capacity to cope with the increased treatment toxicity will be overrepresented in the vaccine group compared with the control group. The lower susceptibility of vaccinated patients remaining at risk can then explain the apparent improved survival in the treatment group (Figure 1C).

#### 4. Eligibility is ascertained using information measured after time zero

To be included in the analyses, patients should have either received a COVID-19 vaccine within 100 days surrounding ICI initiation or should have never received a COVID-19 vaccine throughout follow-up. Thus, patients receiving COVID-19 vaccination within follow-up, but later than 100 days post ICI initiation were excluded from the analyses. This is problematic: the shorter a patient lives, the less likely they are to receive a COVID-19 during follow-up and to be excluded from the analyses. Then, the true survival of patients in the control group is underestimated (Figure 2A). As for immortal-time bias, this survivor-induced selection bias can occur even if the treatment has no effect on survival.

**Figure 2:**
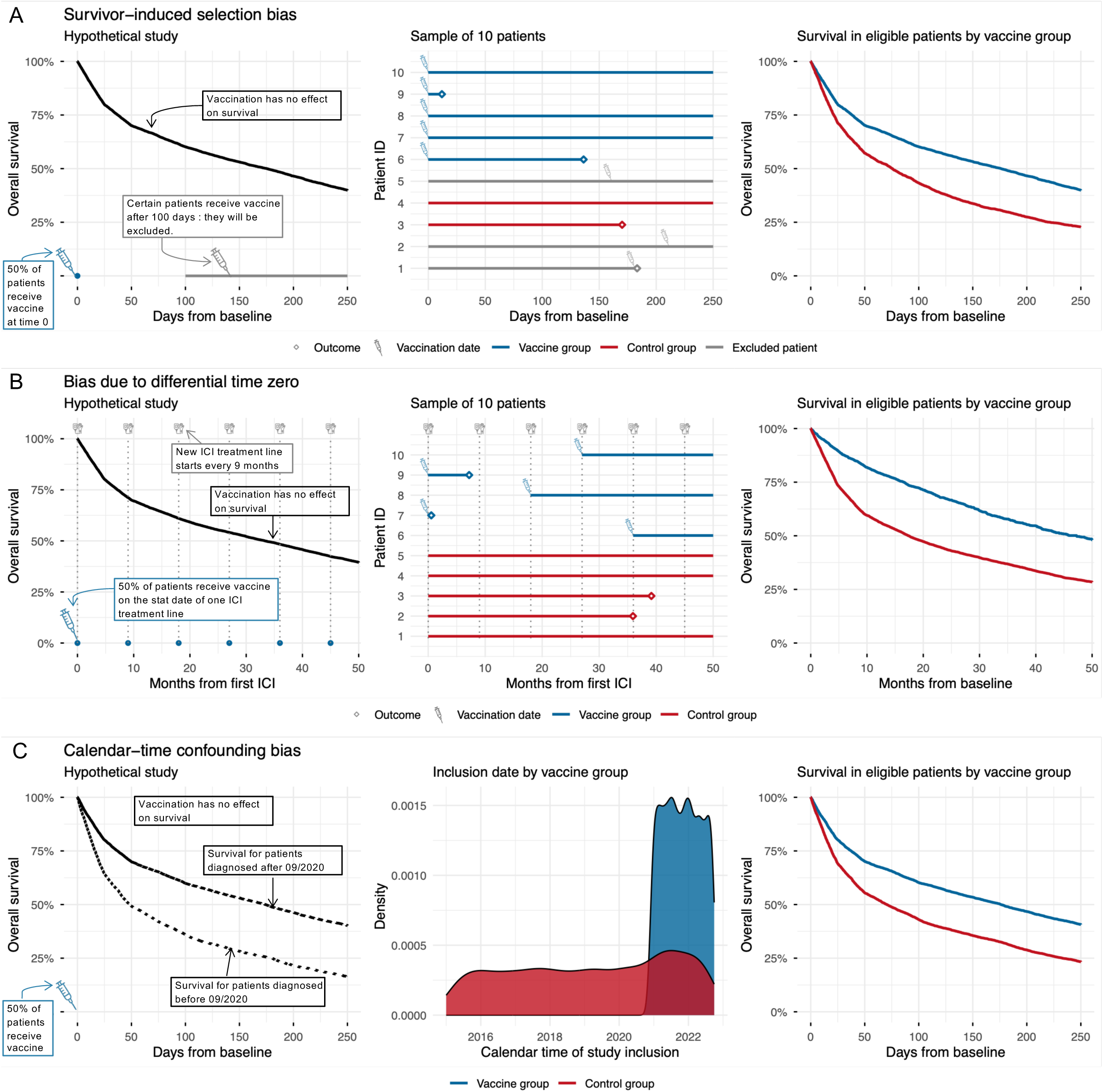
Hypothetical studies illustrating time-related biases 4, 5, and 6 in the context of Grippin et al. (A) Survivor induced selection bias. In this hypothetical study, vaccination has no effect on survival at any time point for any individual. Half of the patients are randomly assigned to receive a vaccine at baseline. Among the other control patients, 30% are randomly selected to receive vaccine at a random time after the first 100 days after baseline. Excluding these patients from the analyses, leads to vaccination appearing spuriously protective. **(B) Bias due to differential definition of time zero in the two intervention groups.** In this hypothetical study, vaccination has no effect on survival at any time point for any individual. Patients start a new ICI treatment line every nine months. Half of patients receive a vaccine on the start date of one of the ICI treatment lines. Setting the time zero for vaccinated units at the start date of the ICI treatment line starting on the day of vaccination leads to vaccination appearing spuriously protective. **(C) Calendar-time confounding bias.** In this hypothetical study, vaccination has no effect on survival at any time point for any individual. Patients can enter the study before September 2020 or after September 2020. Patients entering the study before September 2020 have lower survival probabilities compared to patients entering the study after September 2020, and are all assigned to the control group (historical controls). Among patients entering the study after September 2020, 70% receive a vaccine at study baseline. The presence of calendar-time confounding bias leads to vaccination appearing spuriously protective.

#### 5. Time zero was set differently in the two intervention groups

Time zero was defined differently in the two intervention groups: the unvaccinated individuals were followed from the start date of their first ICI treatment line, while the vaccinated individuals were followed from the start date of the ICI treatment line closest to vaccination. Thus, cancer history at time zero was different between the two groups, which can lead to systematic biases.

As an illustration, suppose that COVID-19 vaccines had no effect on death in the study population. Suppose also that ICI leads to heterogeneous responses in lung cancer patients, with certain patients responding successfully and other not.^19–21^ Then, the survival curve of ICI-treated lung cancer patients declines steeply at the first treatment line because the poor ICI responders die shortly after ICI initiation. Later, the survival curves flatten because the remaining patients are responders whose cancers progress slowly. Postponing time zero at a subsequent ICI treatment line for the vaccinated group thereby overestimates the difference in survival between the two groups (Figure 2B).

#### 6. Widening calendar time of study entry can lead to calendar-time confounding

The authors originally included patients from 2015 onwards, more than five years before the COVID-19 vaccines became available (December 2020). Therefore, patients diagnosed before 2020 were all assigned to the control group. The inclusion of patients diagnosed before 2020 allowed the investigators to increase the sample size of the unvaccinated group, thereby narrowing confidence intervals for survival in the controls. However, the inclusion of such historical controls leads to calendar-time confounding if the risk of experiencing the outcome changes over calendar time (Figure 2C). Differences in the distribution of calendar time entry among treatment and control can also induce administrative censoring to be informative^12^ because control patients are followed longer than treated patients. In particular, the choice of maximum follow-up time matters: with larger follow-up, we need stronger assumptions to ensure that calendar-time biases are avoided (see Gasparollo forthcoming). Overall, there is a bias-variance tradeoff between widening the study years as much as possible to maximize statistical power and inducing calendar-time confounding bias in the study estimates. In Grippin *et al.*, calendar-time confounding is likely a problem because ICI indications changed between 2015 and 2022 to include earlier disease settings.^22^

### Time-related biases can be mitigated by carefully emulating a target trial

Designing a study timeline that avoids time-related biases may appear daunting, but it need not be. A target trial emulation aids with this task. It consists of two steps: (i) describing a hypothetical randomized trial that would answer the research question of interest, and (ii) reproducing its key components as closely as possible using the available observational data. The process of target trial emulation has been extensively discussed before,^2, 3, 3, 10, 12^ and many tutorial and good practices advice are available.^8, 9, 29^ Here, we do not give details, but only briefly illustrate how the study timeline in Grippin *et al.* could have been designed by emulating a hypothetical target trial and comparing the resulting estimates with those reported in the original analysis.

#### Specification of the target trial protocol

We first describe a hypothetical target trial that would answer the study question of Grippin *et al* for the lung cancer cohort. The trial would include patients with stage III/IV NSCLC who initiated ICI treatment as their first systemic treatment after the COVID-19 vaccines became available. Including patients at the first ICI treatment line only ensures that time zero is set at the same disease stage for both intervention groups, thereby avoiding time-related bias 5. Including only patients after the COVID-19 vaccines became available mitigates calendar-time confounding (time-related bias 6). In the target trial, included patients would be randomly assigned to one of the two interventions: (i) get a COVID-19 vaccine in the 100 days following the first ICI or (ii) do not get a COVID-19 vaccine within 100 days of the first ICI. Patients would be followed from the date of randomization (time zero) up to September 2024 (administrative censoring) (Figure 3A-B). The primary outcome would be overall survival (OS) at 36 months. The full protocol of the hypothetical target trial is detailed in Supplementary Table 1. A similar hypothetical target trial for the melanoma cohort of Grippin *et al.* is detailed in Supplementary Table 2.

**Figure 3:**
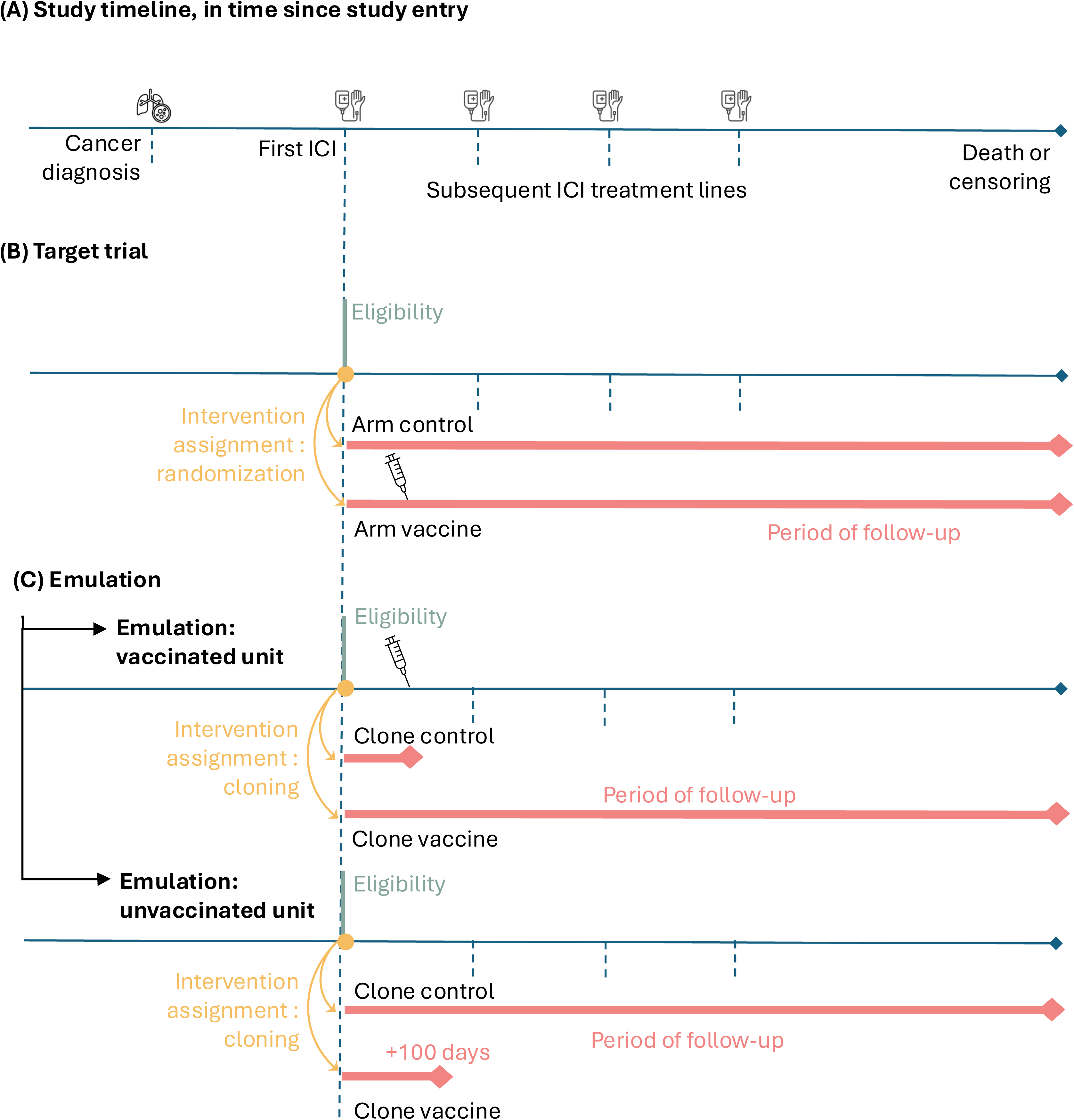
Definition of design-specific time points relative to (A) the study timeline (B) in the hypothetical target trial and (C) in a proposed emulation of the target trial. For the emulation, we distinguish two cases: (i) the patient actually received the vaccine in the 100 days following the start of the first ICI treatment line, (ii) the patient did not receive the vaccine in the 100 days following the start of ICI treatment line. The eligibility flag refers to the start of the eligibility period.

#### Target trial emulation

We emulated the hypothetical target trial using the data presented by Grippin et *al.*, which were available online. Classical difficulties in mimicking the hypothetical trial timeline during the emulation process occur when either (i) the patient meets the eligibility requirement at multiple times of follow-up, or when (ii) treatment strategies are indistinguishable at baseline.^8^ In our example, eligibility is only met once for a patient, at the start date of first ICI treatment line, which also defines time zero. However, patients cannot be assigned to one of the two strategies based on data available at time zero, because of the 100-day grace period authorized for vaccination. Several methods are available to still align the period of intervention assignment with time zero and eligibility in settings with grace period, including the clone-censor-weight and the sequential target trials approaches.^8, 30^ We used a clone-censor-weight approach (Figure 3C).

Given some limitations of the publicly available data, certain time-related biases described previously cannot be avoided. In particular, we could not distinguish patients who received a COVID-19 vaccination in the 100 days before time zero from those receiving COVID-19 vaccination in the 100 data after time zero. Therefore, while the clone-censoring-weight approach mitigates immortal-time bias, our analysis might suffer from prevalent-user bias. <we also did not have access to data on patients who were excluded because they received a COVID-19 vaccination during follow-up, which might lead to an underestimation of survival probabilities in the control group because of survivor-induced selection bias. Further details on the emulation process, including details on covariates used for adjustment, are given in Supplementary Table 1. The code is available online (https://github.com/elisedms/target-trial-emulation-covid-vaccine-ICI).

#### Results

After alignment of time zero at the start date of the first ICI treatment line, exclusion of historical controls, and application of cloning, censoring and weighting to align the period of intervention assignment with time zero and adjusting for confounding, 164 patients were included in the analyses (Supplementary Figure 1). The median age was 65.8 years (IQR 57.7-71.1) and 45.1% of patients were female (Supplementary Table 3). The 3-year OS was estimated to be 45.7% (95% CI 29.6–60.9) in the vaccinated arm and 43.8% (95% CI 28.1–61.1) in the unvaccinated arm (difference: 1.9 percentage-points, 95% CI -23.6–25) (Figure 4). Compared to the original results (Grippin. et al), our analysis of the melanoma cohort also suggests a substantially smaller effect of COVID-19 vaccination on overall survival and progression-free survival (Supplementary Figures 2-3, Supplementary Table 4).

**Figure 4:**
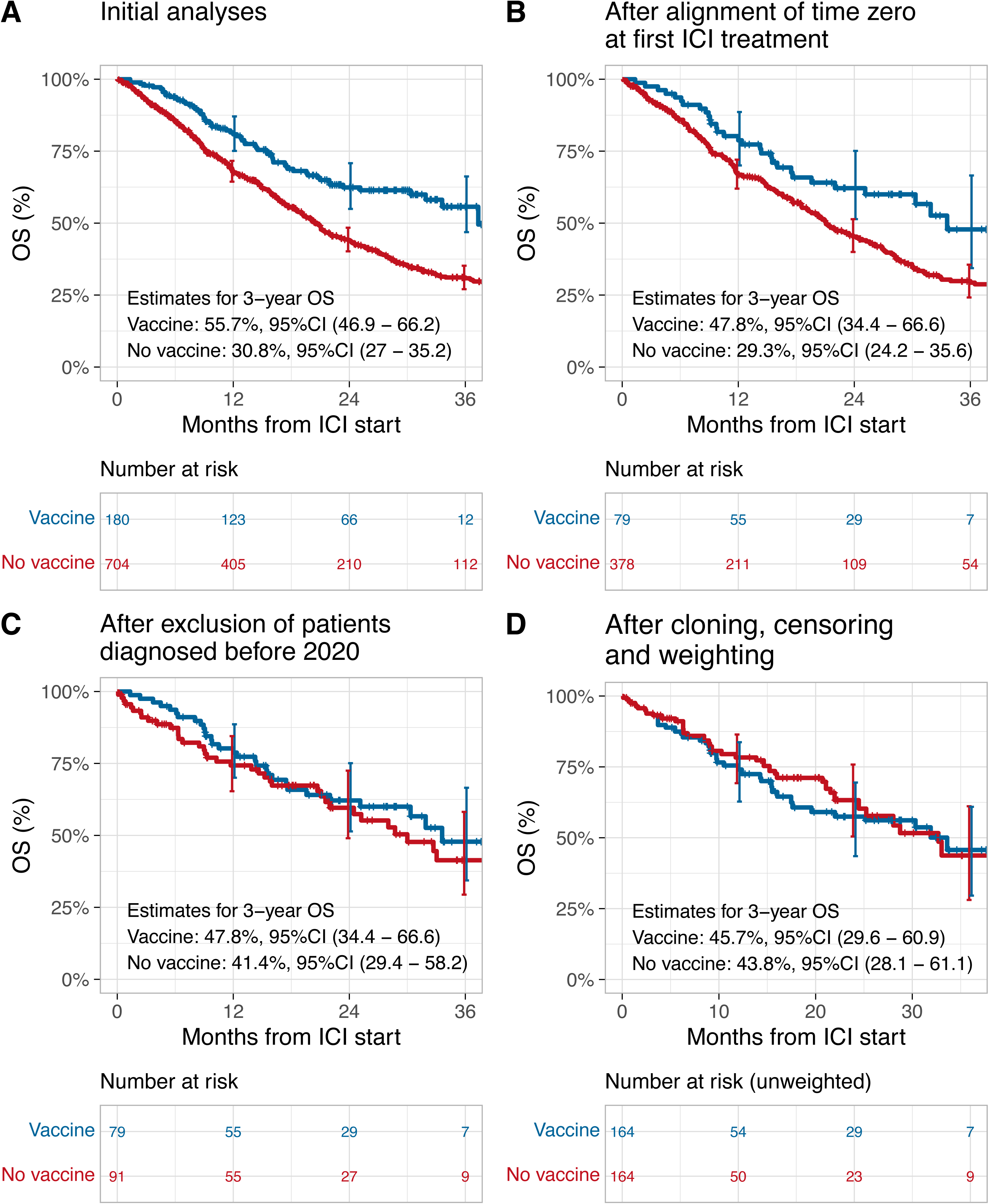
Survival curves, risk tables, and survival estimates after 3 years for the cohort of NSCLC patients. (A) in the original analyses, (B) after alignment of time zero at first ICI treatment by inclusion of patients at ICI initiation as first systemic treatment only, (C) after further exclusion of patients diagnosed before 2020 (historical controls), (D) after further cloning, censoring and weighting. In (D), we also excluded 6 patients with missing ECOG data. In the risk tables of subfigure (D), we present the numbers at risk after cloning but before weighting the patients. The vertical error bars at 12, 24 and 36 months represent confidence intervals around the curves at 1, 2 and 3 years, respectively. Abbreviations: OS: overall survival, ICI: immune checkpoint inhibitors.

## Discussion

Emulating a target trial clarifies the research question by formulating it as a well-defined hypothetical experiment.^2^ This helps to mitigate biases in observational studies, and also to explain why traditional approaches are problematic. Yet, emulating a target trial is not straightforward and requires careful specification and explicit consideration of potential biases. Many such biases are due to events and variables that change over time.

We have discussed these subtle time-related biases and how they can be mitigated and used a published article on COVID-19 vaccines in lung cancer patients (Grippin *et al)* for illustration. We found that the reported protective effect of COVID-19 vaccine on survival outcomes during ICI treatment in Grippin *et al.* could have been driven by multiple time-related biases. In a reanalysis of the study data aiming at avoiding these time-related biases, we did not find evidence of a protective effect of COVID-19 vaccines in ICI-treated lung cancer patients. While addressing key problems of time alignment in the original study, our re-analysis is still prone to time-related biases that cannot be avoided with the data available. Furthermore, we cannot exclude the presence of residual unmeasured confounding bias, for example from time-varying confounders like side effects of ICI treatment, which were not recorded in the data. There could also be bias due to the misspecification of models used for the inverse probability of censoring weights. Finally, the strict eligibility criteria applied to the target trial emulation decreased the effective sample size. Thus, we cannot exclude that there is a (smaller) beneficial effect of COVID-19 vaccines on lung cancer outcomes, as corroborated by biological evidence in Grippin *et al.*, that cannot be detected in the given observational dataset.

## Data Availability

All data can be reproduced using the code available on Github.

## Data availability statement

The data used in this study are publicly available. The source code to reproduce the results is available on Github (https://github.com/elisedms/target-trial-emulation-covid-vaccine-ICI).

## Ethical approval

Ethical approvals for the data used in this study were obtained by the investigators responsible for the original publication. We reanalyzed data that already are publicly available.

## Supplementary material

**Supplementary Table 1:**
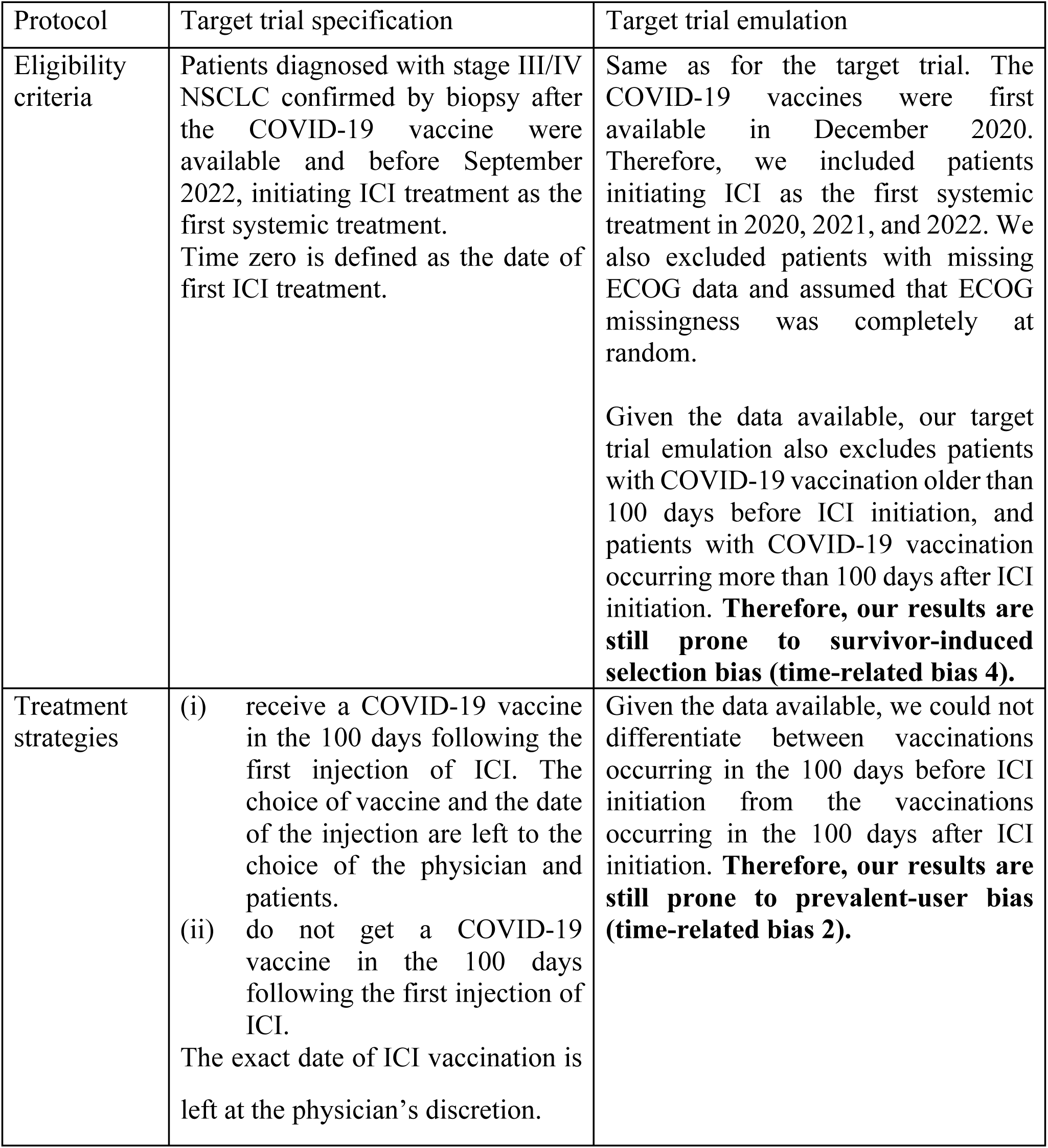
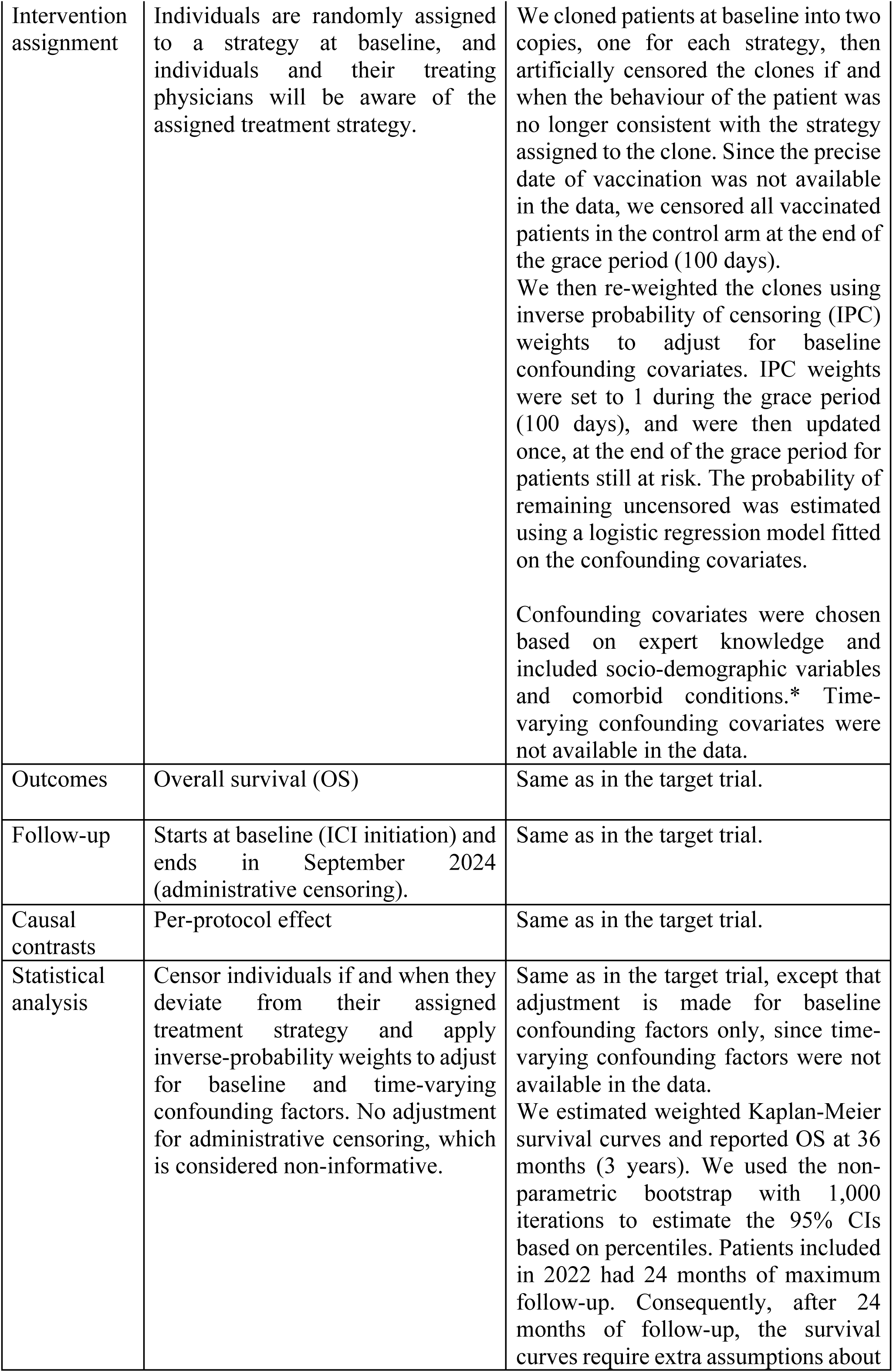
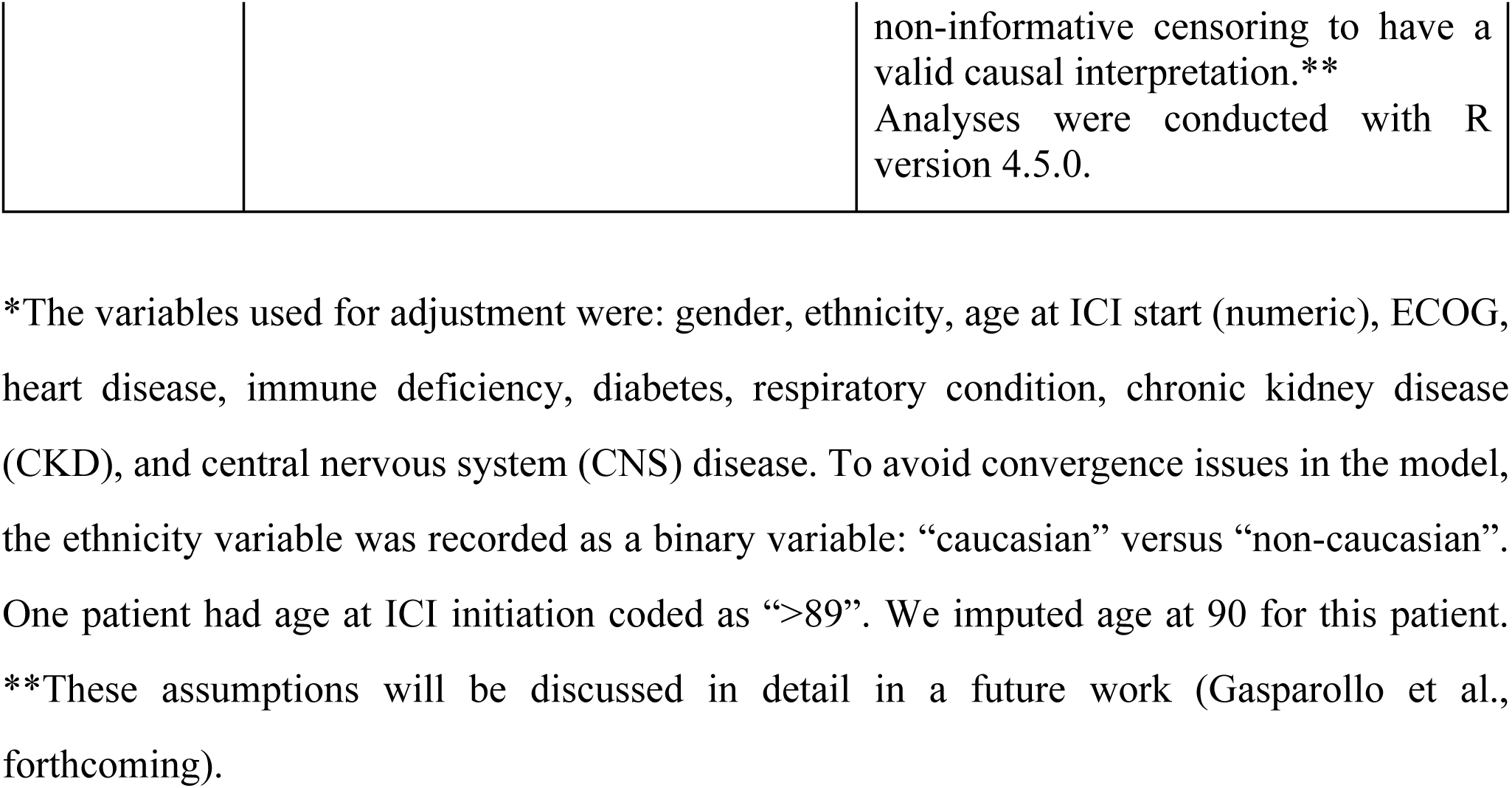
Specification and emulation of a target trial of COVID-19 vaccines during ICI treatment in patients with NSCLC from the data presented in Grippin et al.

| Protocol | Target trial specification | Target trial emulation |
| --- | --- | --- |
| Eligibility criteria | Patients diagnosed with stage III/IV NSCLC confirmed by biopsy after the COVID-19 vaccine were available and before September 2022, initiating ICI treatment as the first systemic treatment.<br>Time zero is defined as the date of first ICI treatment. | Same as for the target trial. The COVID-19 vaccines were first available in December 2020. Therefore, we included patients initiating ICI as the first systemic treatment in 2020, 2021, and 2022. We also excluded patients with missing ECOG data and assumed that ECOG missingness was completely at random.<br>Given the data available, our target trial emulation also excludes patients with COVID-19 vaccination older than 100 days before ICI initiation, and patients with COVID-19 vaccination occurring more than 100 days after ICI initiation. **Therefore, our results are still prone to survivor-induced selection bias (time-related bias 4).** |
| Treatment strategies | (i) receive a COVID-19 vaccine in the 100 days following the first injection of ICI. The choice of vaccine and the date of the injection are left to the choice of the physician and patients.<br>(ii) do not get a COVID-19 vaccine in the 100 days following the first injection of ICI.<br>The exact date of ICI vaccination is left at the physician's discretion. | Given the data available, we could not differentiate between vaccinations occurring in the 100 days before ICI initiation from the vaccinations occurring in the 100 days after ICI initiation. **Therefore, our results are still prone to prevalent-user bias (time-related bias 2).** |
| Intervention assignment | Individuals are randomly assigned to a strategy at baseline, and individuals and their treating physicians will be aware of the assigned treatment strategy. | We cloned patients at baseline into two copies, one for each strategy, then artificially censored the clones if and when the behaviour of the patient was no longer consistent with the strategy assigned to the clone. Since the precise date of vaccination was not available in the data, we censored all vaccinated patients in the control arm at the end of the grace period (100 days).<br>We then re-weighted the clones using inverse probability of censoring (IPC) weights to adjust for baseline confounding covariates. IPC weights were set to 1 during the grace period (100 days), and were then updated once, at the end of the grace period for patients still at risk. The probability of remaining uncensored was estimated using a logistic regression model fitted on the confounding covariates.<br>Confounding covariates were chosen based on expert knowledge and included socio-demographic variables and comorbid conditions.* Time-varying confounding covariates were not available in the data. |
| Outcomes | Overall survival (OS) | Same as in the target trial. |
| Follow-up | Starts at baseline (ICI initiation) and ends in September 2024 (administrative censoring). | Same as in the target trial. |
| Causal contrasts | Per-protocol effect | Same as in the target trial. |
| Statistical analysis | Censor individuals if and when they deviate from their assigned treatment strategy and apply inverse-probability weights to adjust for baseline and time-varying confounding factors. No adjustment for administrative censoring, which is considered non-informative. | Same as in the target trial, except that adjustment is made for baseline confounding factors only, since time-varying confounding factors were not available in the data.<br>We estimated weighted Kaplan-Meier survival curves and reported OS at 36 months (3 years). We used the non-parametric bootstrap with 1,000 iterations to estimate the 95% CIs based on percentiles. Patients included in 2022 had 24 months of maximum follow-up. Consequently, after 24 months of follow-up, the survival curves require extra assumptions about |
|  |  | non-informative censoring to have a valid causal interpretation.**<br>Analyses were conducted with R version 4.5.0. |

\*The variables used for adjustment were: gender, ethnicity, age at ICI start (numeric), ECOG, heart disease, immune deficiency, diabetes, respiratory condition, chronic kidney disease (CKD), and central nervous system (CNS) disease. To avoid convergence issues in the model, the ethnicity variable was recorded as a binary variable: “caucasian” versus “non-caucasian”. One patient had age at ICI initiation coded as “>89”. We imputed age at 90 for this patient. \*\*These assumptions will be discussed in detail in a future work (Gasparollo et al., forthcoming).

**Supplementary Table 2:** Specification and emulation of a target trial of COVID-19 vaccines during ICI treatment in patients with melanoma from the data presented in Grippin et al.

| Protocol | Target trial specification | Target trial emulation |
| --- | --- | --- |
| Eligibility criteria | Patients diagnosed with stage IV melanoma after the COVID-19 vaccines were available and before September 2022, initiating ICI treatment<br>Time zero is defined as the date of first ICI treatment. | Same as for the target trial. The COVID-19 vaccines were first available in December 2020. Therefore, we included patients initiating ICI as the first systemic treatment in 2020, 2021, and 2022.<br>Given the data available, our target trial emulation also excludes patients with COVID-19 vaccination older than 100 days before ICI initiation, and patients with COVID-19 vaccination occurring more than 100 days after ICI initiation. **Therefore, our results are still prone to survivor-induced selection bias (time-related bias 4).** |
| Treatment strategies | (i) receive a COVID-19 vaccine in the 100 days following the first injection of ICI. The choice of vaccine and the date of the injection are left to the choice of the physician and patients.<br>(ii) do not get a COVID-19 vaccine in the 100 days following the first injection of ICI. | Given the data available, we could not differentiate between vaccinations occurring in the 100 days before ICI initiation from the vaccinations occurring in the 100 days after ICI initiation. **Therefore, our results are still prone to prevalent-user bias (time-related bias 2).** |
| Intervention assignment | Individuals are randomly assigned to a strategy at baseline, and individuals and their treating physicians will be aware of the assigned treatment strategy. | We cloned patients at baseline into two copies, one for each strategy, then artificially censored the clones if and when the behaviour of the patient was no longer consistent with the strategy assigned to the clone. Since the precise date of vaccination was not available in the data, we censored all vaccinated patients in the control arm at the end of the grace period (100 days).<br>We then re-weighted the clones using inverse probability of censoring (IPC) weights to adjust for baseline confounding covariates. IPC weights were set to 1 during the grace period (100 days), and were then updated once, at the end of the grace period for patients still at risk. The probability of remaining uncensored was estimated using a logistic regression model fitted on the confounding covariates.<br>Confounding covariates were chosen based on expert knowledge and included socio-demographic variables and comorbid conditions.* Time-varying confounding covariates were not available in the data. |
| Outcomes | Overall survival (OS) and progression-free survival (PFS). | Same as in the target trial. |
| Follow-up | Starts at baseline (ICI initiation) and ends in September 2024 (administrative censoring). | Same as in the target trial. |
| Causal contrasts | Per-protocol effect | Same as in the target trial. |
| Statistical analysis | Censor individuals if and when they deviate from their assigned treatment strategy and apply inverse-probability weights to adjust for baseline and time-varying confounding factors. No adjustment for administrative censoring, which is considered non-informative. | Same as in the target trial, except that adjustment is made for baseline confounding factors only, since time-varying confounding factors were not available in the data.<br>We estimated weighted Kaplan-Meier survival curves and reported OS and PFS at 36 months (3 years). We used the non-parametric bootstrap with 1,000 iterations to estimate the 95% CIs based on percentiles. Patients included in 2022 had 24 months of maximum follow-up. Consequently, after 24 months of follow-up, the |
|  |  | survival curves require extra assumptions about non-informative censoring to have a valid causal interpretation.**<br>Analyses were conducted with R version 4.5.0. |

\*The variables used for adjustment for overall survival were: gender, ethnicity, age at ICI start (numeric), English as primary language, ECOG, heart disease, immune deficiency, diabetes, respiratory condition, chronic kidney disease, and previous history of malignancy at ICI start. The variables used for adjustment for progression-free survival were: gender, age at ICI start (numeric), ECOG, heart disease, immune deficiency, diabetes, respiratory condition, chronic kidney disease, and previous history of malignancy at ICI start. The adjustment sets differed slightly between the two outcomes because we omitted variables leading to convergence issues due to small sample sizes in the logistic model used to estimate the weights. To avoid convergence issues in the model, the ethnicity variable was recorded as a binary variable: “caucasian” versus “non-caucasian”. One patient had age at ICI initiation coded as “>89”. We imputed age at 90 for this patient. \*\*These assumptions will be discussed in detail in a future work (Gasparollo et al., forthcoming).

**Supplementary Table 3:**
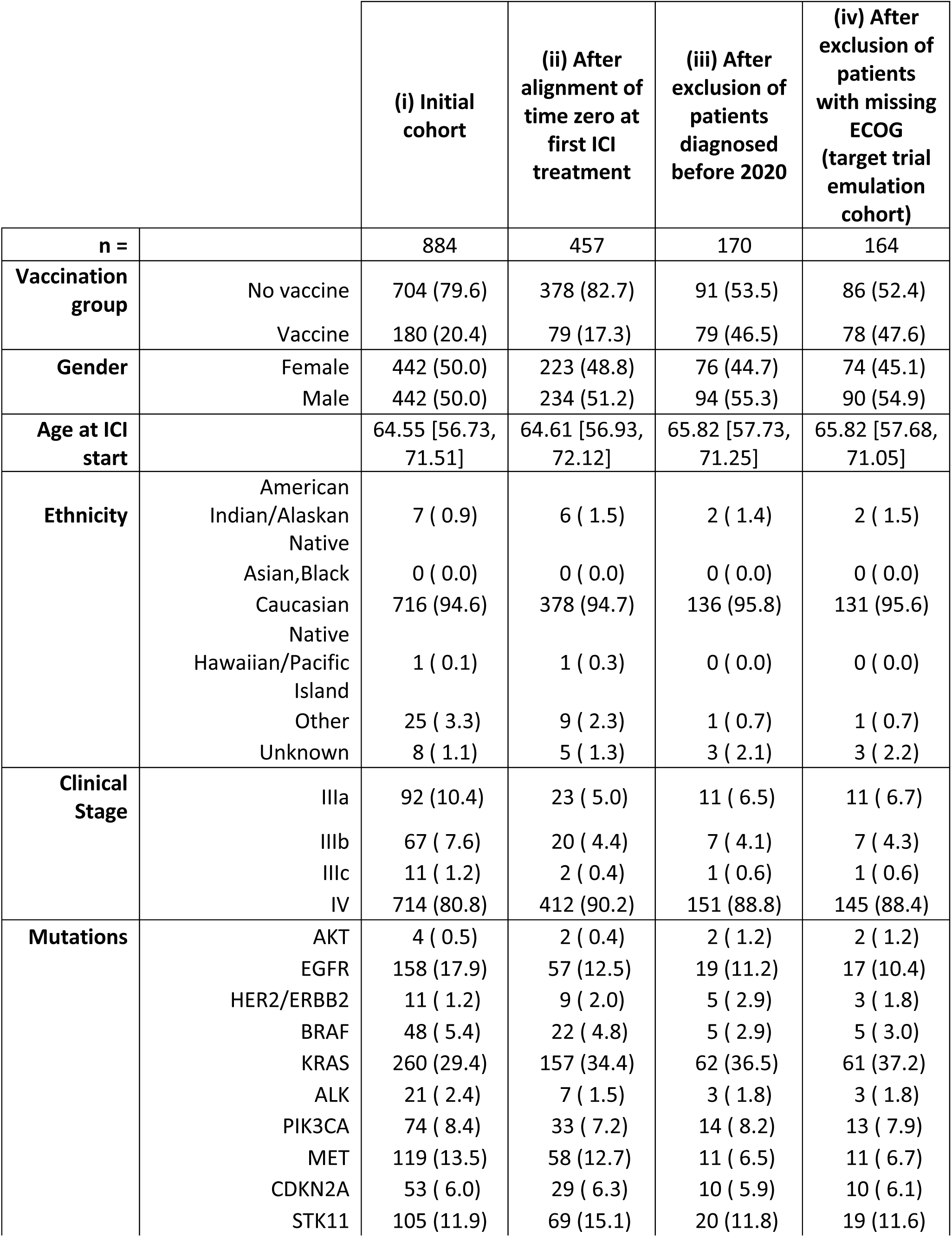
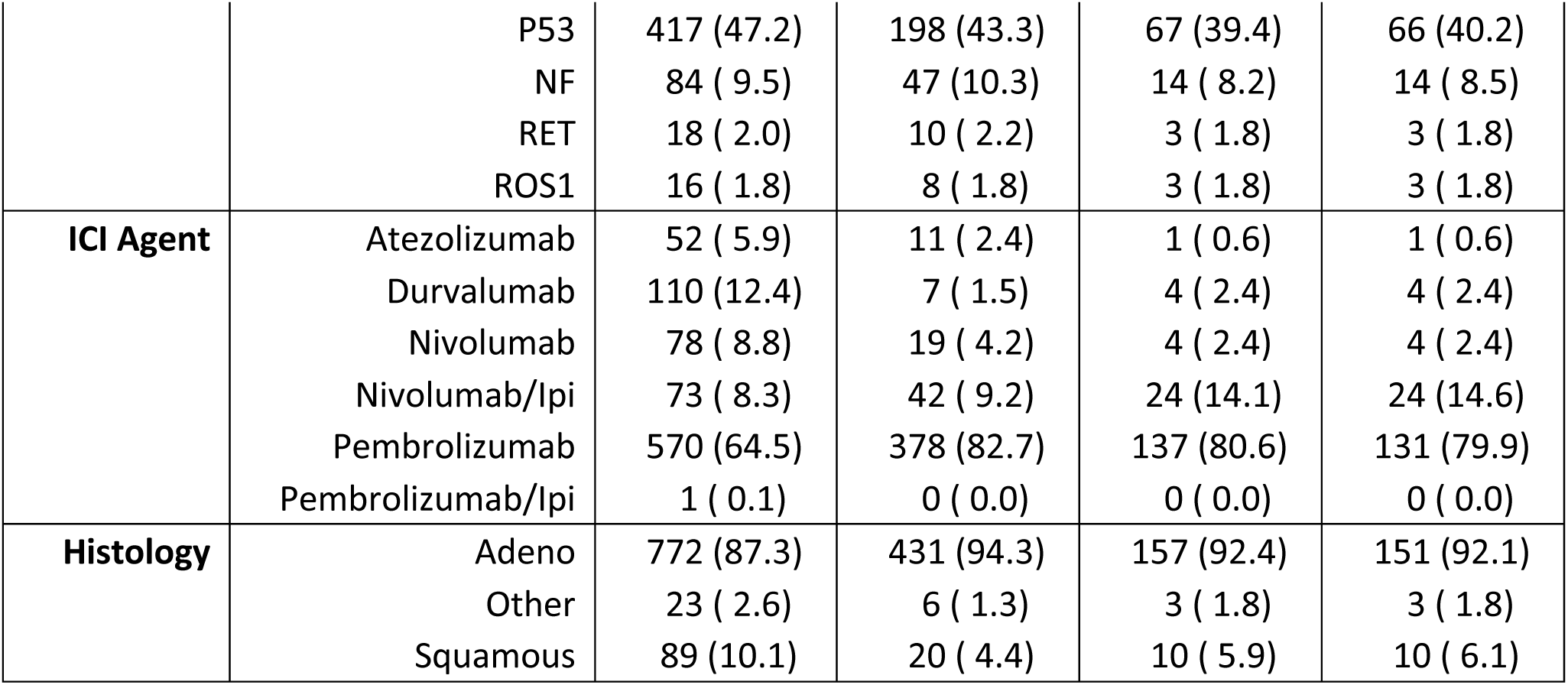
Patient’s characteristics (i) in the initial NSCLC cohort, (ii) after alignment of time zero at first ICI treatment, (iii) after exclusion of patients diagnosed before 2020, and (iv) after exclusion of patients with missing ECOG. Cohort (iv) constitutes the cohort used to emulate the target trial. Abbreviations: ICI: immune checkpoint inhibitors.

|  |  | (i) Initial cohort | (ii) After alignment of time zero at first ICI treatment | (iii) After exclusion of patients diagnosed before 2020 | (iv) After exclusion of patients with missing ECOG (target trial emulation cohort) |
| --- | --- | --- | --- | --- | --- |
| **n =** |  | 884 | 457 | 170 | 164 |
| **Vaccination group** | No vaccine | 704 (79.6) | 378 (82.7) | 91 (53.5) | 86 (52.4) |
|  | Vaccine | 180 (20.4) | 79 (17.3) | 79 (46.5) | 78 (47.6) |
| **Gender** | Female | 442 (50.0) | 223 (48.8) | 76 (44.7) | 74 (45.1) |
|  | Male | 442 (50.0) | 234 (51.2) | 94 (55.3) | 90 (54.9) |
| **Age at ICI start** |  | 64.55 [56.73, 71.51] | 64.61 [56.93, 72.12] | 65.82 [57.73, 71.25] | 65.82 [57.68, 71.05] |
| **Ethnicity** | American Indian/Alaskan Native | 7 ( 0.9) | 6 ( 1.5) | 2 ( 1.4) | 2 ( 1.5) |
|  | Asian,Black | 0 ( 0.0) | 0 ( 0.0) | 0 ( 0.0) | 0 ( 0.0) |
|  | Caucasian | 716 (94.6) | 378 (94.7) | 136 (95.8) | 131 (95.6) |
|  | Native Hawaiian/Pacific Island | 1 ( 0.1) | 1 ( 0.3) | 0 ( 0.0) | 0 ( 0.0) |
|  | Other | 25 ( 3.3) | 9 ( 2.3) | 1 ( 0.7) | 1 ( 0.7) |
|  | Unknown | 8 ( 1.1) | 5 ( 1.3) | 3 ( 2.1) | 3 ( 2.2) |
| **Clinical Stage** | IIIa | 92 (10.4) | 23 ( 5.0) | 11 ( 6.5) | 11 ( 6.7) |
|  | IIIb | 67 ( 7.6) | 20 ( 4.4) | 7 ( 4.1) | 7 ( 4.3) |
|  | IIIc | 11 ( 1.2) | 2 ( 0.4) | 1 ( 0.6) | 1 ( 0.6) |
|  | IV | 714 (80.8) | 412 (90.2) | 151 (88.8) | 145 (88.4) |
| **Mutations** | AKT | 4 ( 0.5) | 2 ( 0.4) | 2 ( 1.2) | 2 ( 1.2) |
|  | EGFR | 158 (17.9) | 57 (12.5) | 19 (11.2) | 17 (10.4) |
|  | HER2/ERBB2 | 11 ( 1.2) | 9 ( 2.0) | 5 ( 2.9) | 3 ( 1.8) |
|  | BRAF | 48 ( 5.4) | 22 ( 4.8) | 5 ( 2.9) | 5 ( 3.0) |
|  | KRAS | 260 (29.4) | 157 (34.4) | 62 (36.5) | 61 (37.2) |
|  | ALK | 21 ( 2.4) | 7 ( 1.5) | 3 ( 1.8) | 3 ( 1.8) |
|  | PIK3CA | 74 ( 8.4) | 33 ( 7.2) | 14 ( 8.2) | 13 ( 7.9) |
|  | MET | 119 (13.5) | 58 (12.7) | 11 ( 6.5) | 11 ( 6.7) |
|  | CDKN2A | 53 ( 6.0) | 29 ( 6.3) | 10 ( 5.9) | 10 ( 6.1) |
|  | STK11 | 105 (11.9) | 69 (15.1) | 20 (11.8) | 19 (11.6) |
|  | P53 | 417 (47.2) | 198 (43.3) | 67 (39.4) | 66 (40.2) |
|  | NF | 84 ( 9.5) | 47 (10.3) | 14 ( 8.2) | 14 ( 8.5) |
|  | RET | 18 ( 2.0) | 10 ( 2.2) | 3 ( 1.8) | 3 ( 1.8) |
|  | ROS1 | 16 ( 1.8) | 8 ( 1.8) | 3 ( 1.8) | 3 ( 1.8) |
| **ICI Agent** | Atezolizumab | 52 ( 5.9) | 11 ( 2.4) | 1 ( 0.6) | 1 ( 0.6) |
|  | Durvalumab | 110 (12.4) | 7 ( 1.5) | 4 ( 2.4) | 4 ( 2.4) |
|  | Nivolumab | 78 ( 8.8) | 19 ( 4.2) | 4 ( 2.4) | 4 ( 2.4) |
|  | Nivolumab/Ipi | 73 ( 8.3) | 42 ( 9.2) | 24 (14.1) | 24 (14.6) |
|  | Pembrolizumab | 570 (64.5) | 378 (82.7) | 137 (80.6) | 131 (79.9) |
|  | Pembrolizumab/Ipi | 1 ( 0.1) | 0 ( 0.0) | 0 ( 0.0) | 0 ( 0.0) |
| **Histology** | Adeno | 772 (87.3) | 431 (94.3) | 157 (92.4) | 151 (92.1) |
|  | Other | 23 ( 2.6) | 6 ( 1.3) | 3 ( 1.8) | 3 ( 1.8) |
|  | Squamous | 89 (10.1) | 20 ( 4.4) | 10 ( 5.9) | 10 ( 6.1) |

**Supplementary Table 4:** Patient’s characteristics (i) in the initial melanoma cohort and (ii) after exclusion of patients diagnosed before 2020. Cohort (ii) constitutes the cohort used to emulate the target trial. Abbreviations: ICI: immune checkpoint.

|  |  | (i) Initial cohort | (ii) After exclusion of patients diagnosed before 2020 (target trial emulation cohort) |
| --- | --- | --- | --- |
| **n =** |  | 210 | 134 |
| **Vaccination group** | Observed no vaccine | 167 (79.5) | 91 (67.9) |
|  | Observed vaccine | 43 (20.5) | 43 (32.1) |
| **Gender** | Female | 71 (33.8) | 50 (37.3) |
|  | Male | 139 (66.2) | 84 (62.7) |
| **Age at ICI start** |  | 64.33 [53.25, 73.60] | 63.87 [50.80, 74.28] |
| **Ethnicity** | Asian | 2 ( 1.0) | 2 ( 1.5) |
|  | Black | 3 ( 1.4) | 1 ( 0.7) |
|  | Hispanic | 9 ( 4.3) | 6 ( 4.5) |
|  | Other | 1 ( 0.5) | 1 ( 0.7) |
|  | Unknown | 6 ( 2.9) | 2 ( 1.5) |
|  | White | 189 (90.0) | 122 (91.0) |
| **Mutations** | NRAS | 43 (20.5) | 23 (17.2) |
|  | BRAF | 79 (37.6) | 46 (34.3) |
|  | KIT | 20 ( 9.5) | 11 ( 8.2) |
|  | PTEN | 18 ( 8.6) | 10 ( 7.5) |
|  | GNA11 | 4 ( 1.9) | 4 ( 3.0) |
|  | GNAQ | 3 ( 1.4) | 1 ( 0.7) |
| **ICI drug** | Avelumab | 1 ( 0.5) | 0 ( 0.0) |
|  | Ipilimumab | 1 ( 0.5) | 0 ( 0.0) |
|  | Ipilimumab/Nivolumab | 99 (47.1) | 67 (50.0) |
|  | Nivolumab | 75 (35.7) | 43 (32.1) |
|  | Pembrolizumab | 28 (13.3) | 19 (14.2) |
|  | Pembrolizumab/Ipi | 6 ( 2.9) | 5 ( 3.7) |

**Supplementary Figure 1:**
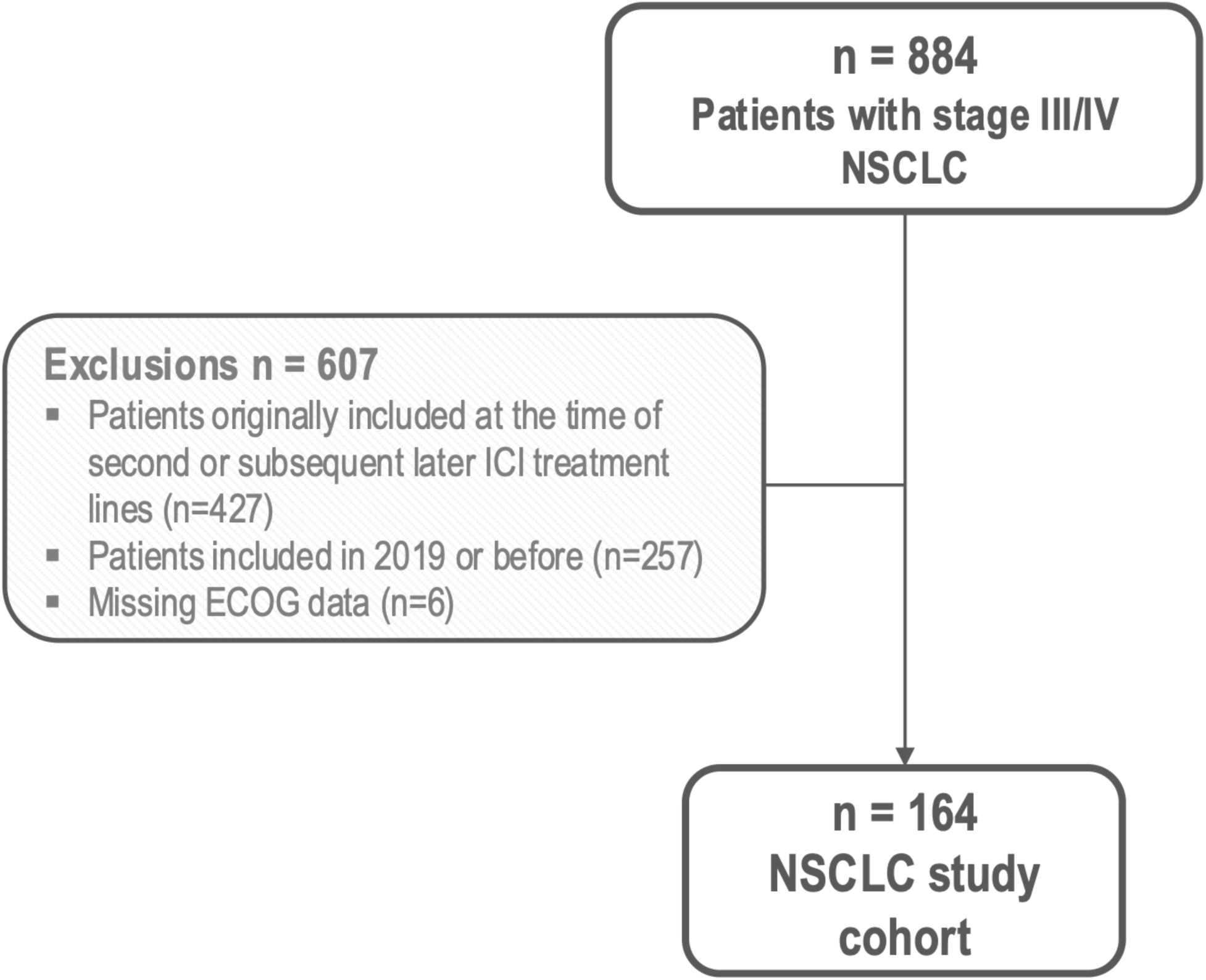
Study flowchart for the NSCLC cohort and for (B) the stage IV melanoma cohort.

**Supplementary Figure 2:**
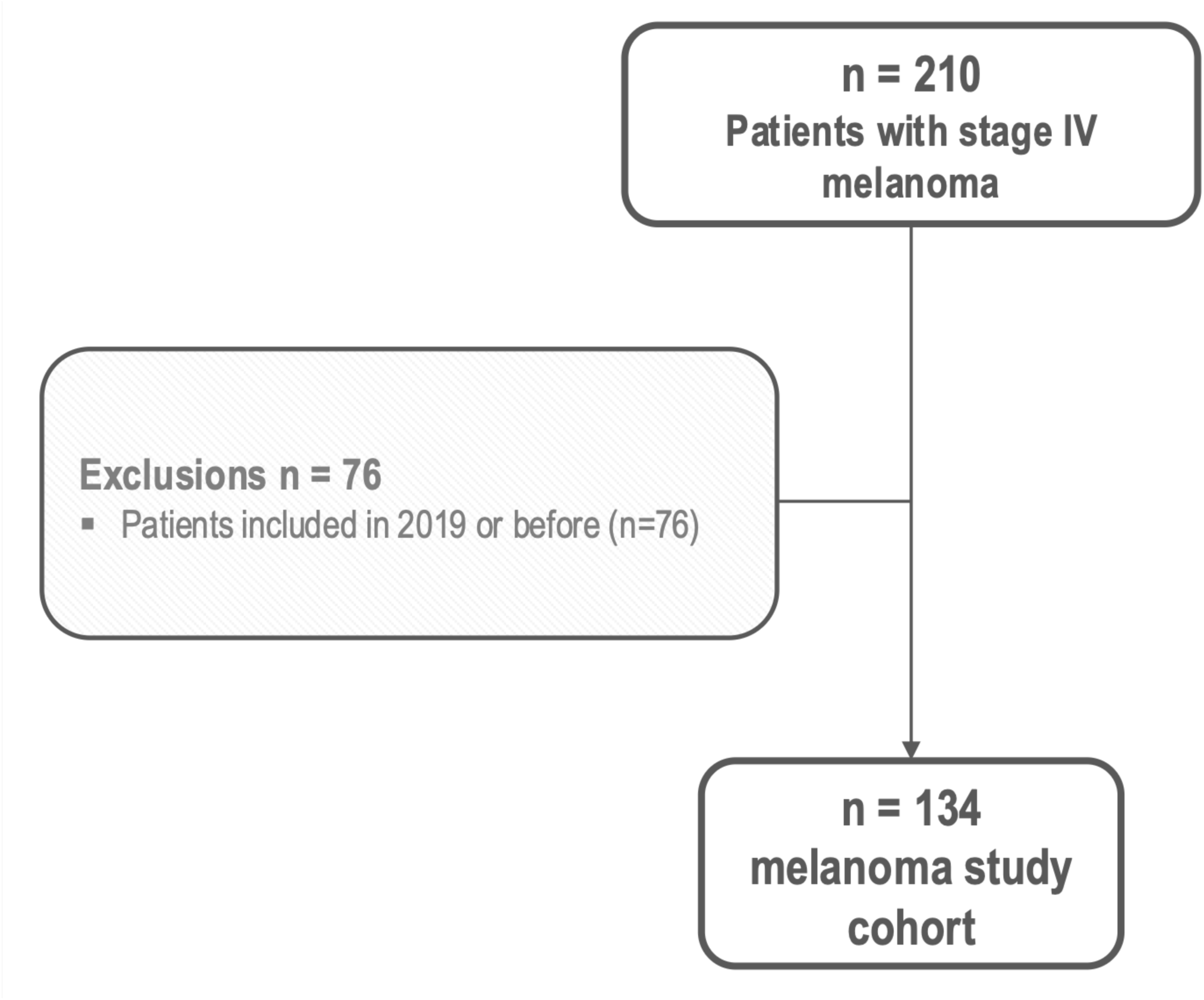
Study flowchart for the stage IV melanoma cohort.

**Supplementary Figure 3:**
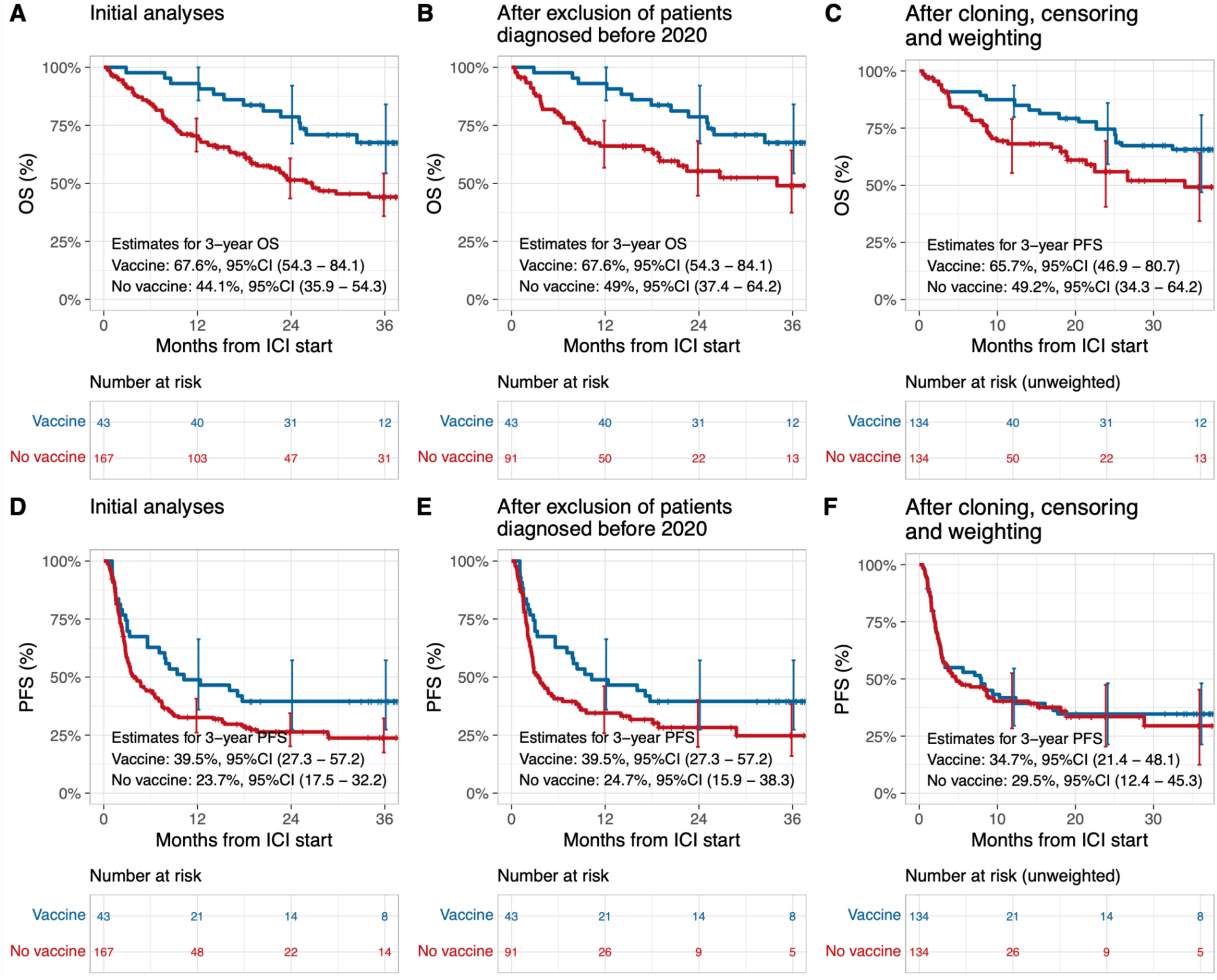
Survival curves, risk tables, and OS and PFS estimates after 3 years for the cohort of melanoma patients. (A) in the original analyses for overall survival, (B) after further exclusion of patients diagnosed before 2020 (historical controls) for overall survival, (C) after further cloning, censoring and weighting for overall survival, (D) in the original analyses for progression-free survival, (E) after further exclusion of patients diagnosed before 2020 (historical controls) for progression-free survival, (F) after further cloning, censoring and weighting for progression-free survival. In the risk tables of subfigures (C) and (F), we present the numbers at risk after cloning but before weighting the patients. The vertical error bars at 12, 24 and 36 months represent confidence intervals around the curves at 1, 2 and 3 years, respectively. Abbreviations: OS: overall survival, PFS: progression-free survival, ICI: immune checkpoint inhibitors.

